# Uptake and timing of viral load testing and frequency of viraemic episodes during pregnancy in South Africa

**DOI:** 10.1101/2025.03.19.25324178

**Authors:** Nelly Jinga, Karl-Günter Technau, Kate Clouse, Nkosinathi Ngcobo, Cornelius Nattey, Candice Hwang, Anna Grimsrud, Amy Wise, Nicola van Dongen, Thalia Ferreira, Maanda Mudau, Mhairi Maskew

**Author notes:** **Corresponding author:** Nelly Jinga Health Economics and Epidemiology Research Office, Building C, First Floor, Sunnyside Office Park, 32 Princess of Wales Terrace, Parktown, Johannesburg, 2193.

## Abstract

**Background:** Repeated monitoring of viral load (VL) among pregnant women living with HIV (WLWH) is critical in vertical transmission prevention. For women who are newly diagnosed with HIV during pregnancy, a subsequent VL is recommended three months after ART initiation, and for all women living with HIV, follow-up VL is required every six months throughout pregnancy and breastfeeding [2]. Here, we describe the uptake and timing of VL testing and frequency and distribution of viraemic episodes during pregnancy.

**Methods:** We linked prospective cohort data from WLWH whose infants were born at Rahima Moosa Mother and Child Hospital (RMMCH) in Johannesburg, South Africa (2013-2018) to laboratory data from the National Health Laboratory Services national HIV cohort. We report the uptake and timing of VL testing, and frequency of viremia and viral suppression. We also explore factors associated with having at least one or more VL test.

**Results:** Data from 4,064 women with known dates of entry into antenatal care and delivery during the study period were analysed. Overall, less than half (46%) completed VL testing during pregnancy. Most VL were conducted during the third trimester (67%). Only 5% (n=100) were during the first trimester and 11% within 7 days of delivery. Three-quarters of tests during pregnancy indicated viral suppression (VL <400 copies/mL), 7% viraemic (VL 400-1000 copies/mL), and 19% high grade viraemia (VL >1000 copies/mL). We found that being older (≥35) and being engaged in HIV care prior to pregnancy were significantly associated with VL testing during pregnancy.

**Conclusion:** With less than half of pregnant women living with HIV in this study having a VL measure during their pregnancy, and VL testing occurring late in pregnancy, this study highlights critical gaps in providing quality HIV care to women and prevention of vertical transmission.

## BACKGROUND

For pregnant women living with HIV, repeated monitoring of viral load (VL) is critical in vertical transmission prevention; every week a pregnant woman maintains an elevated viral load heightens the risk of vertical transmission [1]. Progress towards elimination of vertical transmission requires sustained maternal VL suppression from conception through pregnancy to the postpartum period at the end of breastfeeding. Given the narrow window of opportunity for intervention in case of an elevated maternal VL there is need to implement regular maternal VL monitoring and respond quickly to elevated VL.

For women who are newly diagnosed with HIV during pregnancy, a subsequent VL is recommended three months after ART initiation, and for all women living with HIV, follow-up VL is required every six months throughout pregnancy and breastfeeding [2]. South Africa adopted the Option B+ policy in 2015, which expanded lifelong antiretroviral therapy (ART) to pregnant women living with HIV, and expanded again to universal access in 2016. Maternal viral load testing guidelines recommended testing at pregnancy diagnosis, and VL was recommended at first antenatal care presentation regardless of when the last VL was done [2]. In 2019, the guidelines were updated to include an additional VL test at delivery and three-monthly during breastfeeding [1]. Prior to Option B+ (between 2013 and 2015); the WHO-recommended Option B approach was adopted in 2013, and recommended women to do a VL test at pregnancy confirmation at 6 and 12 months post-initiation.

In order to monitor treatment effectiveness and ensure the prevention of vertical transmission of HIV, timely VL testing is essential in the care of pregnant women living with HIV. VL monitoring enables clinicians to detect early virologic failure (HIV RNA≥1,000 copies/mL) and take the necessary action. In South Africa, entry to antenatal care (ANC) tends to occur later than the recommended first 12 weeks of gestation [3][4] with the median gestational age at entry to ANC ranging from 14 to 23 weeks [6–10]. Gestational age at ANC entry influences the timing of VL testing during pregnancy, especially for newly-diagnosed and/or ART-naïve pregnant women yet this relationship is not well described in relation to policies of expanded access to ART.

The NHLS National HIV Cohort includes the laboratory data of nearly all patients receiving HIV care in the public sector in South Africa since 2004. Compared to clinical cohorts, the NHLS National HIV Cohort offers benefits. The data are extracted from patient charts into databases straight from the source testing platforms, making them less prone to data input errors and the cohort represents trends of treatment-seeking in the public sector since the data include all lab-monitored HIV patients receiving care in that sector [6].

Here, we linked an established maternal cohort to routinely collected laboratory records and describe the frequency and temporal distribution of VL testing and viraemic episodes among pregnant WLWH attending ANC in Johannesburg, South Africa.

## METHODS

### Study design and data sources

Rahima Moosa Mother and Child Hospital (RMMCH) is a large academic obstetrics, gynaecology and paediatric referral centre and functions as the primary delivery facility of a sub-district of Johannesburg, South Africa [7]. The Department of Paediatrics and Child Health offers both in-hospital admissions as well as outpatient facilities. RMMCH provides care to patients from its immediate catchment area but also acts as a regional referral centre for high risk or complex neonatal, paediatric and obstetrics/gynaecology cases. There were 13,072 infants delivered in 2018 at RMMCH.

The Rahima Moosa Maternal cohort collected maternal and infant data from approximately 10,000 WLWH 18 years and older attending ANC at RMMCH between 2013 and 2018, and captured detailed clinical information, including ANC booking date, delivery date, HIV testing dates, early infant diagnosis testing, prophylaxis and maternal blood results relevant to HIV captured at the point of delivery. Approximately 18% of women delivering at RMMCH are living with HIV [8].

The National Health Laboratory Service (NHLS) provides diagnostic laboratory and pathology services to approximately 80% of the South African population via public-sector facilities. Previously, graph-based linkage techniques have been applied to HIV monitoring laboratory records from the NHLS to create the NHLS National HIV Cohort; a longitudinal cohort of people living with HIV accessing care in South Africa’s public sector [6]. The cohort included the laboratory data of nearly all persons receiving HIV care in the public sector since April 2004. Individuals were included in the cohort if their laboratory records include at least one CD4 count or HIV RNA VL test. Data collected include age, sex, facility location and test results for HIV-associated monitoring and care.

### Data linkage and validation

The NHLS centrally allocates alphanumeric barcodes for all biological samples submitted to its laboratories. All tests conducted on an individual biological specimen are identified through these unique barcodes in both the NHLS and RMMCH cohorts. For this analysis, we applied deterministic record linkage procedures to link maternal records from the RMMCH cohort to the NHLS National HIV Cohort using test sample barcodes. The barcode linkage procedure was validated through a random sample (n=1200) of records using patient surname, first name, and date of birth. In this manually matched set of RMMCH records, 96% were confirmed matches in the NHLS cohort; and the remaining 4% of records were not linked to any NHLS data barcode. All women who booked for ANC at RMMCH and had records linked to the NHLS National HIV Cohort were included in the analysis.

### Study Variables

We considered the pregnancy period as the period between recorded date of first ANC visit and date of delivery. We report three primary outcomes related to VL testing during pregnancy. First, <u>uptake of VL testing</u> was defined according to VL test records in either the NHLS cohort or RMMCH cohort datasets during the pregnancy period and was classified into three categories: 1) no VL test observed, 2) one VL test observed and 3) more than one VL test observed. Second, we classified <u>timing of VL testing during pregnancy</u> according to dates of VL test results recorded in the NHLS or RMMCH cohort datasets and categorized timing of test as either: first trimester (gestation: weeks 1 to 13 weeks, 6 days), second trimester (gestation: weeks 14 weeks to 27 weeks, 6 days) or third trimester (gestation: weeks 28 to delivery), using trimester definitions established by the American College of Obstetrics and Gynaecologists [9]. Our third outcome was <u>VL test result</u>. For this outcome, each VL test result was classified into one of three categories: 1) suppressed VL (VL <400 copies/mL); 2) low grade viraemia (VL 400-1000 copies/mL) or 3) high grade viraemia (VL >1000 copies/mL). The VL test outcome was estimated in two ways: first at earliest test during pregnancy (Outcome 3i) and then at test within 90 days of entry to antenatal care (Outcome 3ii). We also assessed if changes in the South African national ART policy impacted observed maternal VL testing. National policy eras were categorised as “Prior to Option B+” for the period between January 2013 and December 2015, while the “Option B+ era” is defined as the period from January 2015 to dataset censor date (April 2018). Lastly, we explored factors associated with having at least one VL test using log binomial regression to estimate the relative risk (RR) and corresponding 95% confidence interval.

### Statistical analysis

The characteristics of pregnant women included in the analysis at time of delivery were described using frequencies and simple proportions. We then summarised the frequency of VL testing between first ANC visit and delivery stratified by gestational age at VL test. Frequency of VL outcomes during pregnancy were described with simple frequencies. Next, we summarised the VL result categories by gestational age at test and time to first observed VL during pregnancy with medians and report interquartile ranges. We report two estimates for viral load results: 1) a “population level” viral load which aggregates viral load results among all women in the cohort regardless of whether a VL test was observed or not; and 2) VL results restricted to those with an observed viral load test. For the log binomial regression, crude relative risks (RR) and adjusted relative risks (aRR) with 95% confidence intervals (CI) relative risk were reported. The change in estimate (CIE) approach was applied to identify the factors to include in the final model. Predictors were retained in the final model if their addition altered the estimate of the primary exposure by more than 10%, indicating a meaningful contribution to the model.

### Ethical considerations

This study was approved by the Human Research Ethics Committee (HREC) of the University of Witwatersrand and Boston University. This is a secondary analysis of existing data collected as part of routine care. Permission for anonymised patient laboratory records in NHLS was obtained from the NHLS. Informed consent was obtained from patients in RMMCH cohort.

## RESULTS

Of 4,671 women who had their records linked to the NHLS HIV cohort, 40 (<1%) were missing a date of delivery and 565 (12%) had VL tests observed outside the pregnancy period; these were excluded from the final analysis. We analysed data on 4,064 mothers with known dates of entry into antenatal care and delivery during the study period, the analysis was limited to VL during pregnancy and delivery. Characteristics of women at entry to antenatal care are summarised in **Table 1**. The median maternal age was 30 years (IQR:26-34); median gestation at birth was 39 weeks (IQR: 38-40); median antenatal CD4 was 394 cells/mm^3^ (IQR :248-554); median parity was 2 (IQR: 2-3); median gravidity was 3 (IQR: 2-3) and 80% of the women were on a standard first line regimen of tenofovir, emtricitabine and efavirenz (TDF/FTC/EFV) [10].

**Table 1.** Characteristics of women included in the analysis (n=4,064).

| Characteristic |  | N (%) |
| --- | --- | --- |
| Age at delivery | Median (IQR) | 30 (26-34) |
|  | 15-19 | 78 (1.9) |
|  | 20-24 | 664 (16.3) |
|  | 25-29 | 1174 (28.9) |
|  | 30-34 | 1252 (30.8) |
|  | 35-39 | 703 (17.3) |
|  | 40-44 | 181 (4.5) |
|  | 45+ | 12 (0.3) |
| Parity | Median (IQR) | 2 (2-3) |
|  | 1 | 626 (15.4) |
|  | 2 | 1614 (39.7) |
|  | 3 | 1177 (29.0) |
|  | 4 | 459 (11.3) |
|  | 5-9 | 179 (4.4) |
|  | Missing | 9 (0.2) |
| Gravidity | Median (IQR) | 3 (2-3) |
|  | 1 | 467 (11.5) |
|  | 2 | 1387 (34.1) |
|  | 3 | 1286 (31.6) |
|  | 4 | 606 (14.9) |
|  | 5-10 | 309 (7.6) |
|  | Missing | 9 (0.2) |
| Gestational age at birth (weeks) | Median (IQR) | 39 (38-40) |
| Gestational age at entry into antenatal care (weeks) | Median (IQR) | 20 (15-26) |
| Timing of entry into antenatal care | Trimester 1 | 758 (18.7) |
|  | Trimester 2 | 2480 (61.3) |
|  | Trimester 3 | 808 (20.0) |
| **Antenatal CD4 (cells/μL)** | Median (IQR) | 394 (248-554) |
| **Regimen at entry into antenatal care among those on ART** | FDC (TDF/FTC/EFV) | 3250 (80.0) |
|  | TDF/3TC/EFV | 254 (6.3) |
|  | TDF/3TC/Aluvia | 30 (0.7) |
|  | TDF/3TC/NVP | 96 (2.4) |
|  | AZT/3TC/EFV | 40 (1.0) |
|  | Other combinations | 276 (6.8) |
|  | Missing | 118 (2.9) |

### Timing of entry into antenatal care

Median gestation at entry into ANC was 20 weeks (IQR:15-26). Overall, 19% (758) of women first attended ANC in the first trimester, 61% (2480) during the second trimester and 20% (808) during the third trimester (Table 1). The proportion of women presenting for ANC in trimester 3 dropped from 69% in 2014 to 60% in 2015 (**Supplementary Figure 1**).

### Uptake and timing of VL testing

Overall, 2,184 (54%) women had no VL observed at any time between ANC booking and delivery, while 1,880 (46%) women had at least one VL observed during the same period (**Table 2**). Two-thirds (67%) of VL testing occurred during the third trimester with a peak at 34 weeks’ gestation **(Figure 1)**. Although 19% of women attended antenatal care in the first trimester, only 5% of VL tests (n=100) occurred during the first trimester. Delivery testing was low: 11% had a VL within 7 days of delivery and 69 (4%) completed a VL at delivery. The median gestational age at first VL test for women who initiated ART during ANC was higher than for women who initiated ART before ANC: 34 weeks (IQR: 29-37) vs. 28 weeks (IQR: 21-35) **(Table 2)**. There were no significant changes in the proportion of women getting their first VL in each trimester over the period 2014 to 2017 **(Supplementary Figure 2)**.

**Figure 1.**
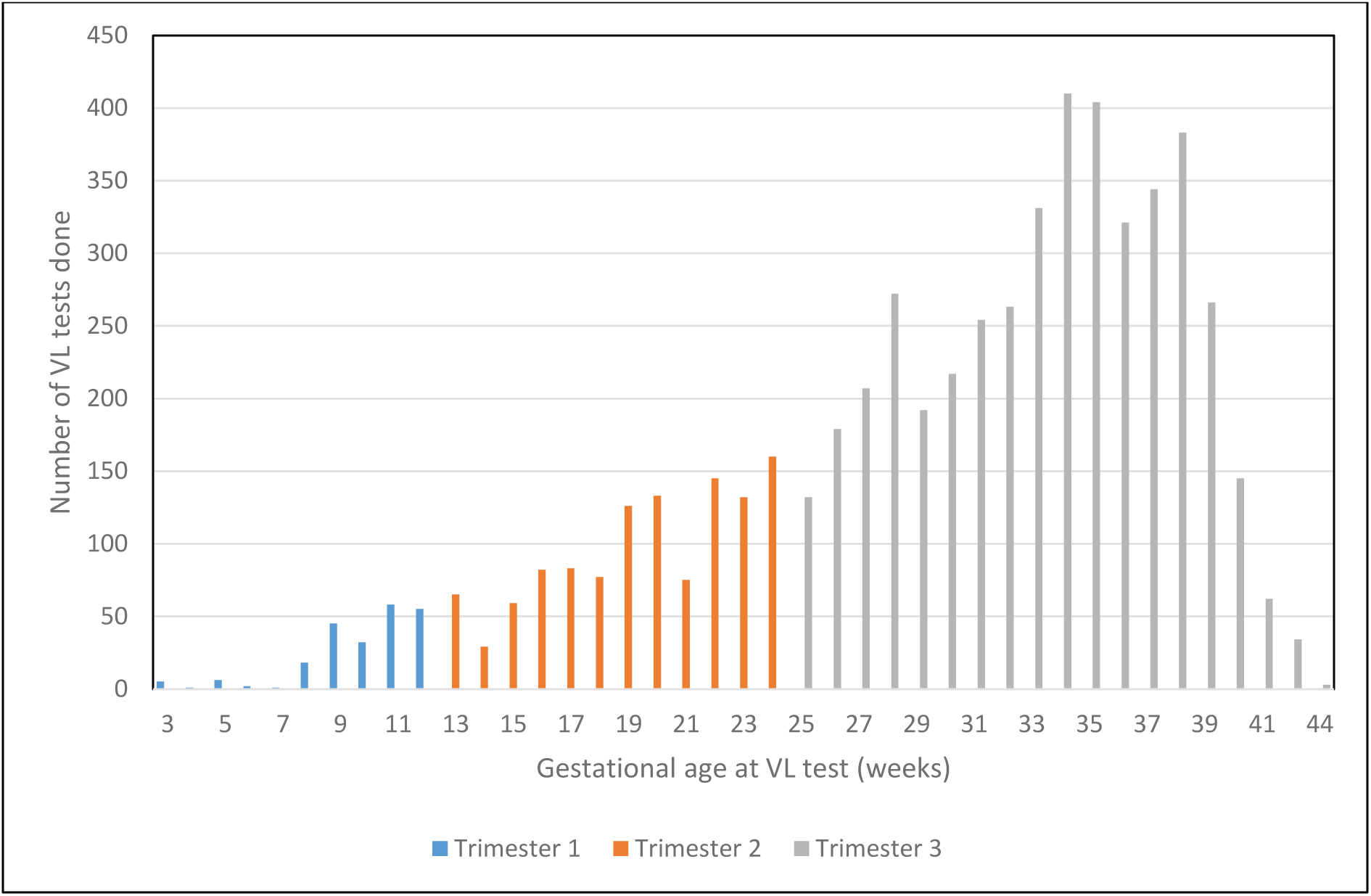
Frequency and timing of VL testing during pregnancy.

**Table 2.** Uptake of viral load testing guidelines during the study period (2013-2018).

|  | N (%) |
| --- | --- |
| **Median gestational presentation (IQR)** | 20 (15-26) |
| **% on ART and presentation to antenatal care** | 3977 (98%) |
| **Median time between ANC booking and observed VL (days) (IQR)** | 1 (1-51) |
| **Median time from ANC booking to first suppressed VL (days)** | 43 (1-107) |
| **Uptake of observed VL monitoring** | N=4064 |
| No VL observed during pregnancy | 2177 (54%) |
| At least 1 VL observed during pregnancy | 1869 (46%) |
| *Only 1 test* | 1742 (43%) |
| *More than 1 test* | 127 (3%) |
| **Timing of observed VL monitoring** | n=1869 |
| Trimester 1 | 100 (5%) |
| Trimester 2 | 563 (30%) |
| Trimester 3 | 1206 (65%) |
| Within 7 days of delivery | 212 (11%) |
| On the day of delivery | 69 (4%) |
| Median gestational age at test (initiated ART before entry to ANC) | 28 (21-35) |
| Median gestational age at test (initiated ART during ANC) | 34 (29-37) |
| **VL result during pregnancy (VL for the first test)** | n=1869 |
| Suppressed <400 copies/mL | 1376 (74%) |
| Viraemic episode 400-1000 copies/mL | 132 (7%) |
| Unsuppressed >1000 copies/mL | 361 (19%) |
| **VL result at entry to ANC** | n=1073 |
| Suppressed <400 copies/mL | 730 (68%) |
| Viraemic episode 400-1000 copies/mL | 89 (8%) |
| Unsuppressed >1000 copies/mL | 254 (24%) |

The proportion of women with no VL testing observed decreased over the study period 2014-2017 (87% in 2014 and 52% in 2017), though uptake was still insufficient to meet program targets by the end of the study period at 68% (**Figure 2**).

**Figure 2.**
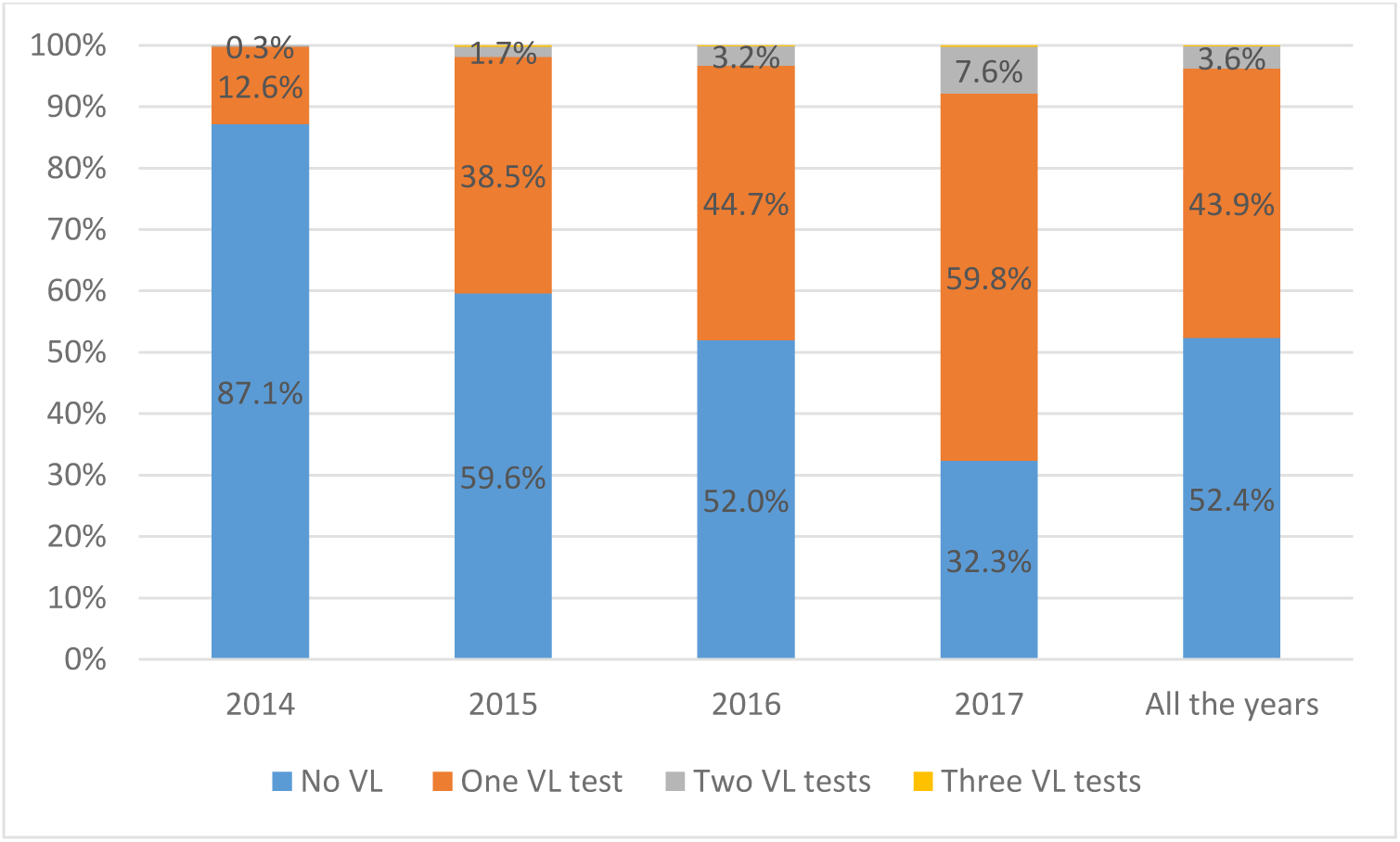
Viral load suppression category stratified by gestational age at test.

### VL results during pregnancy

Overall, three-quarters of VL tests observed during pregnancy indicated viral suppression, 7% showed low grade viraemia, and 20% high grade viraemia. At the population level, this translates to overall VL suppression rate of 34% during pregnancy.

**Figure 3** summarizes the distribution of the first VL result by gestational age after entry into ANC during the pregnancy period. The proportion of high grade viraemia varied by gestational age, peaking at nearly 30% at around 17 weeks’ gestation. The proportion of women with first suppressed VL test during pregnancy increased from 61% in trimester 3 in 2014 to 73% in trimester 3 in 2017 (**Supplementary Figure 3**).

**Figure 3.**
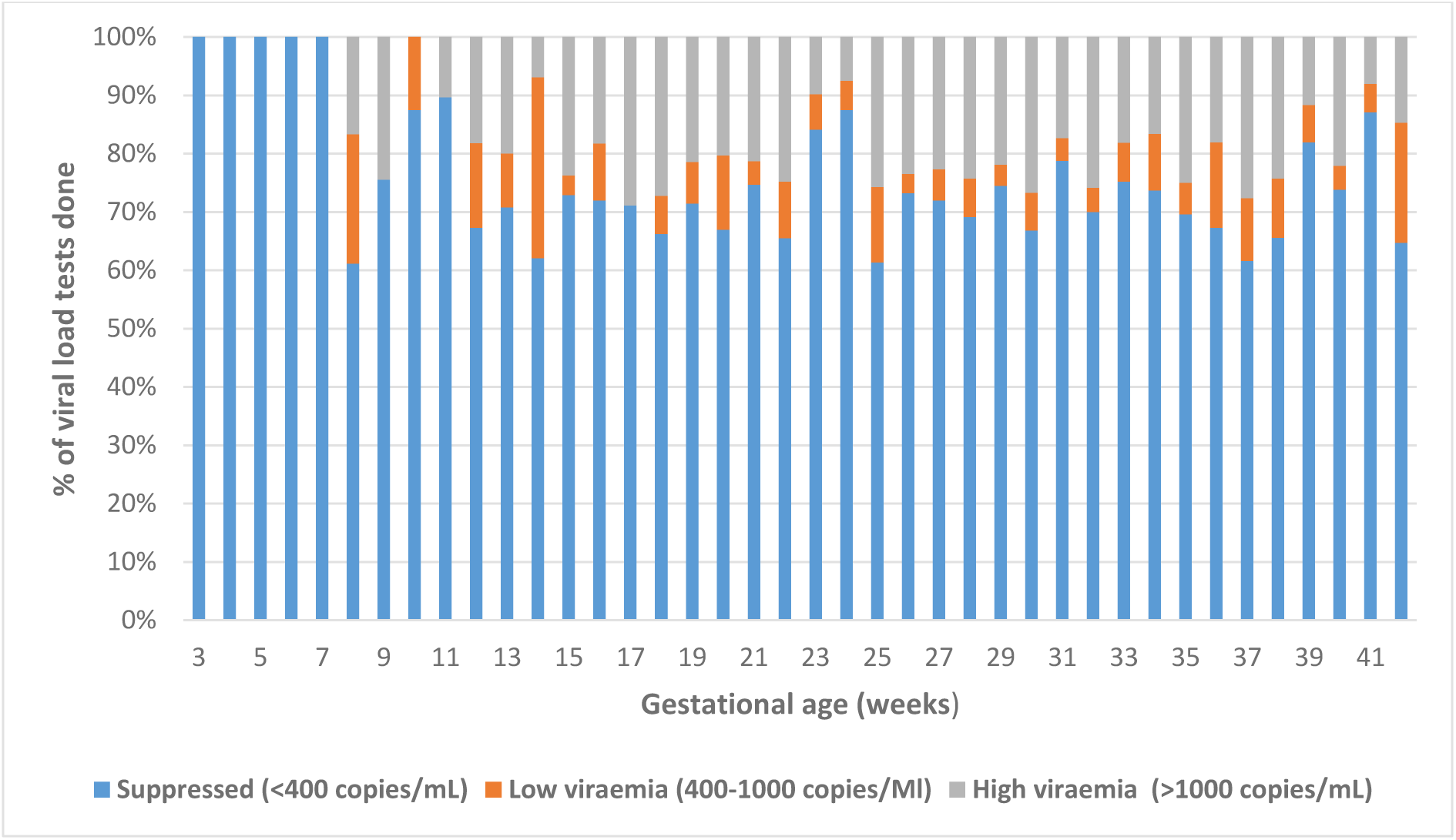
Viral load suppression category stratified by gestational age at test.

### Factors associated with having a VL test during pregnancy

The frequency of VL testing also improved with increasing maternal age (**Figure 4**). Compared with younger women, women aged 35-44 were almost 20% more likely to have at least one VL during pregnancy (aRR=1.2, 95% CI: 1.1-1.3) **(Table 3)**. Evidence of prior engagement in care was significantly associated with having a VL test during pregnancy (aRR=1.6, 95% CI: 1.5-1.9). Parity, Gravidity, timing of entry into ANC, gestation at entry into ANC and year of VL testing were not associated with having a VL test. (Table 3).

**Figure 4.**
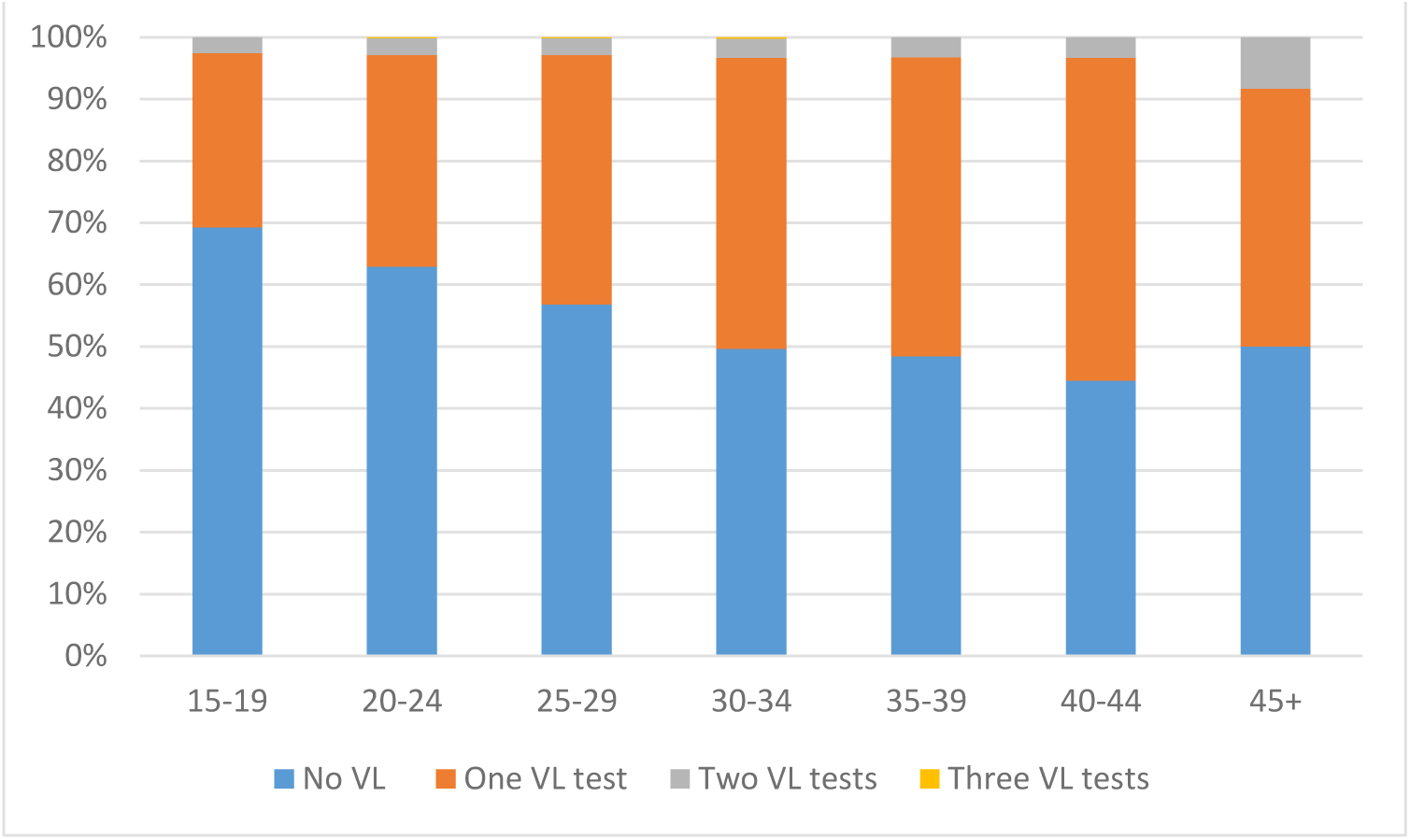
Uptake of VL testing by age group.

**Table 3.** Factors associated with having at least one VL test during pregnancy.

|  | N=1869 | Crude OR (95% CI) | Adjusted OR (95% CI) |
| --- | --- | --- | --- |
| **Maternal age at delivery** |  |  |  |
| 15-24 | 270 (14%) | ref | ref |
| 25-34 | 1133 (60%) | 1.3 (1.16-1.42) | 1.2 (1.05-1.40) |
| 35-44 | 460 (25%) | 1.4 (1.28-1.61) | 1.2 (1.06-1.47) |
| 45+ | 6 (0%) | 1.3 (0.77-2.43) | 0.7 (0.22-2.34) |
| **Parity** |  |  |  |
| Low multi-parity (1-3) | 1834 (98%) | ref |  |
| Grand multipara (4-9) | 338 (18%) | 1.01 (0.92-1.10) |  |
| **Gravidity** |  |  |  |
| 1 | 192 (10%) | ref |  |
| 2 | 611 (33%) | 1.1 (0.91-1.26) |  |
| 3 | 614 (33%) | 1.2 (0.99-1.37) |  |
| 4 | 294 (16%) | 1.2 (0.99-1.42) |  |
| 5-10 | 153 (8%) | 1.2 (0.99-1.50) |  |
| **First VL test year** |  |  |  |
| 2014 | 46 (2%) | ref |  |
| 2015 | 520 (28%) | 3.1 (2.34-4.09) | 2.8 (4.81-16.37) |
| 2016 | 657 (35%) | 3.7 (2.80-4.87) | 11.65 (5.53-24.55) |
| 2017 | 646 (35%) | 5.14 (3.9-6.76) | 19.1 (8.30-43.79) |
| **Engagement in care** |  |  |  |
| No prior engagement in care | 459 (25%) | ref | ref |
| Evidence of prior engagement in care | 1393 (75%) | 1.8 (1.66-1.97) | 2.6 (2.20-2.96) |
| **Timing of observed VL monitoring** |  |  |  |
| First trimester | 468 (25%) | ref | ref |
| Second trimester | 1177 (63%) | 0.8 (0.71-0.82) | 0.64 (0.52-0.78) |
| Third trimester | 224 (12%) | 0.4 (0.39-0.51) | 0.30 (0.22-0.39) |
| Booking Year |  |  |  |
| 2014 | 91 (5%) | ref | ref |
| 2015 | 530 (28%) | 1.8 (1.51-2.22) | 0.6 (0.32-1.03) |
| 2016 | 714 (38%) | 2.3 (1.90-2.77) | 0.6 (0.44-0.96) |
| 2017 | 534 (29%) | 3.1 (2.53-3.70) |  |
| **Gestation at entry into ANC** |  |  |  |
| <median gestation of 21 weeks | 1106 (59%) | ref | ref |
| >=median gestation of 21 weeks | 763 (41%) | 0.7 (0.63-0.72) | 0.7 (0.57-0.82) |

### Impact of Option B+ policy implementation on VL testing

The occurrence of an observed VL during pregnancy (adjusted for CD4 at entry into ANC, gestational age at delivery and maternal age at entry into ANC in the second trimester), was three times more likely during Option B+ compared to prior to Option B+ (aRR=3.04, 95% CI: 2.34-3.94). The proportion of women with no VL observed decreased during the Option B+ era, compared to before the policy implementation (aRR=0.69, 95% CI: 0.62-0.78). Compared to the pre-Option B+ era, the completion of VL testing improved more than two-fold during the Option B+ era (20% vs 49%, respectively).

## DISCUSSION

Despite the importance of routine and timely monitoring of VL during pregnancy, in this analysis of the completion and frequency of VL testing among pregnant women in South Africa from 2013-2018, we found several key areas of concern and opportunities for action. First, less than half of women studied had a VL measure during their pregnancy. Additionally, overall VL suppression during pregnancy was 34%, far below suppression rates of 74% observed in the 2019 South African Antenatal HIV Sentinel Survey of 8,112 pregnant women aged 15 to 49 [11]. Although our results suggest an improvement in uptake of VL testing over the years and across policies, our findings indicate that considerable work is still needed to ensure VL monitoring is aligned with normative guidance and best practice. Monitoring of VL remains a critical gap in prevention of vertical transmission of HIV and optimising HIV care among pregnant women living with HIV. The low rates of VL testing coverage among pregnant women in our study, though worrying, is in keeping with other estimates of VL testing coverage during pregnancy. Other reports of VL testing indicate low coverage among pregnant women with estimates ranging from 30% in South Africa [12] and 32% in Zimbabwe [13]. In Kenya, 50% of pregnant women had a VL test during ANC while 28% of women living with HIV did not have any documented VL testing by six months postpartum [14].

Second, we noted that the majority of VL testing occurred late in pregnancy. Not surprisingly, women who initiated ART during ANC were more likely to have first VL test during the third trimester compared to women who started ART before entry to ANC. This increase in testing in the third trimester could be attributed to newly HIV-diagnosed women who only completed a first VL three months after entry into ANC, or due to women presenting late for ANC [15], [16], or both. Presenting late for ANC delays not only VL monitoring during pregnancy, but also interventions such as enhanced adherence counselling and regimen switching in case of treatment failure. Only 7% of VL tests were done within seven days of delivery since at this time, the guidelines had not been updated to include VL testing at delivery [1].

The low coverage of VL testing and lack of adherence to guidelines during pregnancy is a cause for concern and future work should explore the specific barriers and challenges that pregnant women and or clinics face in adhering to viral load testing guidelines. Low access and uptake of VL testing during pregnancy could be addressed by implementing innovative approaches like point-of-care viral load testing during ANC [17].

Finally, our results highlighted that younger mothers remain vulnerable to poor treatment outcomes during pregnancy. Our results showed that younger women were less likely to have a VL test during pregnancy compared to older women. Younger women are less likely to have a VL during pregnancy since they are less likely to be on ART [18][19]. It is important for policies and programmes to have interventions that target younger women. Our findings also show that being engaged in HIV care prior to pregnancy improves the timely VL testing thereby improving prevention of vertical transmission.

Our results should be interpreted in light of some limitations of our study. First, the study period data was limited to the period 2013-2018 which did not allow us to evaluate trends in maternal VL testing as universal access to ART was widely implemented in South Africa. Second, as this cohort represents a single facility, results may not be generalised more broadly to other settings in South Africa. Despite these limitations, this analysis was strengthened through linkage of maternal records to the NHLS National HIV cohort, a longitudinal cohort observing laboratory testing associated with HIV monitoring at the national level and robust to silent transfers. This improved ascertainment of VL testing outcomes beyond the facility-level observations available in maternal records [6][20].

### Conclusion

With less than half of pregnant women living with HIV in this study having a VL measure during their pregnancy, this study highlights a critical gap in providing quality prevention of vertical transmission and HIV care to women. VL monitoring is a critical tool for assessing maternal and infant outcomes, but frequency has been alarmingly low throughout our study period. Coverage of VL testing remained incomplete and testing patterns do not always reflect what the guidelines recommend. The three-month lag in VL testing for newly diagnosed women results in the bulk of viral load testing occurring in the third trimester with low uptake at delivery and inadequate viral suppression rates. Additional years of data will give information on how the changes in the guidelines at delivery impacted VL testing patterns.

## Ethics approval and consent to participate

Approval for this study was obtained from the University of the Witwatersrand, Human Research Ethics Committee (Wits HREC) (protocol M200237, approved 15 June 2020). This is a secondary analysis of existing data collected as part of routine care. Permission for anonymised patient laboratory records in NHLS was obtained from the NHLS. Informed consent was obtained from patients in RMMCH cohort.

## Consent for publication

Not applicable.

## Availability of data and materials

The data that support the findings of this study are owned by the NHLS and Rahima Moosa Mother and Child Hospital, and access is governed by policies and procedures in response to requests made directly to the Department and the hospital. As such, the study teams will not have authority to release the data to the public or other data-sharing repositories. However, these data can be requested by the public through standardized request forms, which are then considered in an internal review procedure.

## Authors’ contributions

MM*, NJ and K-G T conceptualized the study and methodology. NJ and MM* performed the analysis. NJ wrote the original draft. MM** and KC provided interpretation of results NN, CN, CH, AG, AW, NVD, and TF critically reviewed the manuscript.

*MM* - Mhairi Maskew*

*MM** - Maanda Mudau*

## Supplementary Figures

**Supplementary Figure 1.**
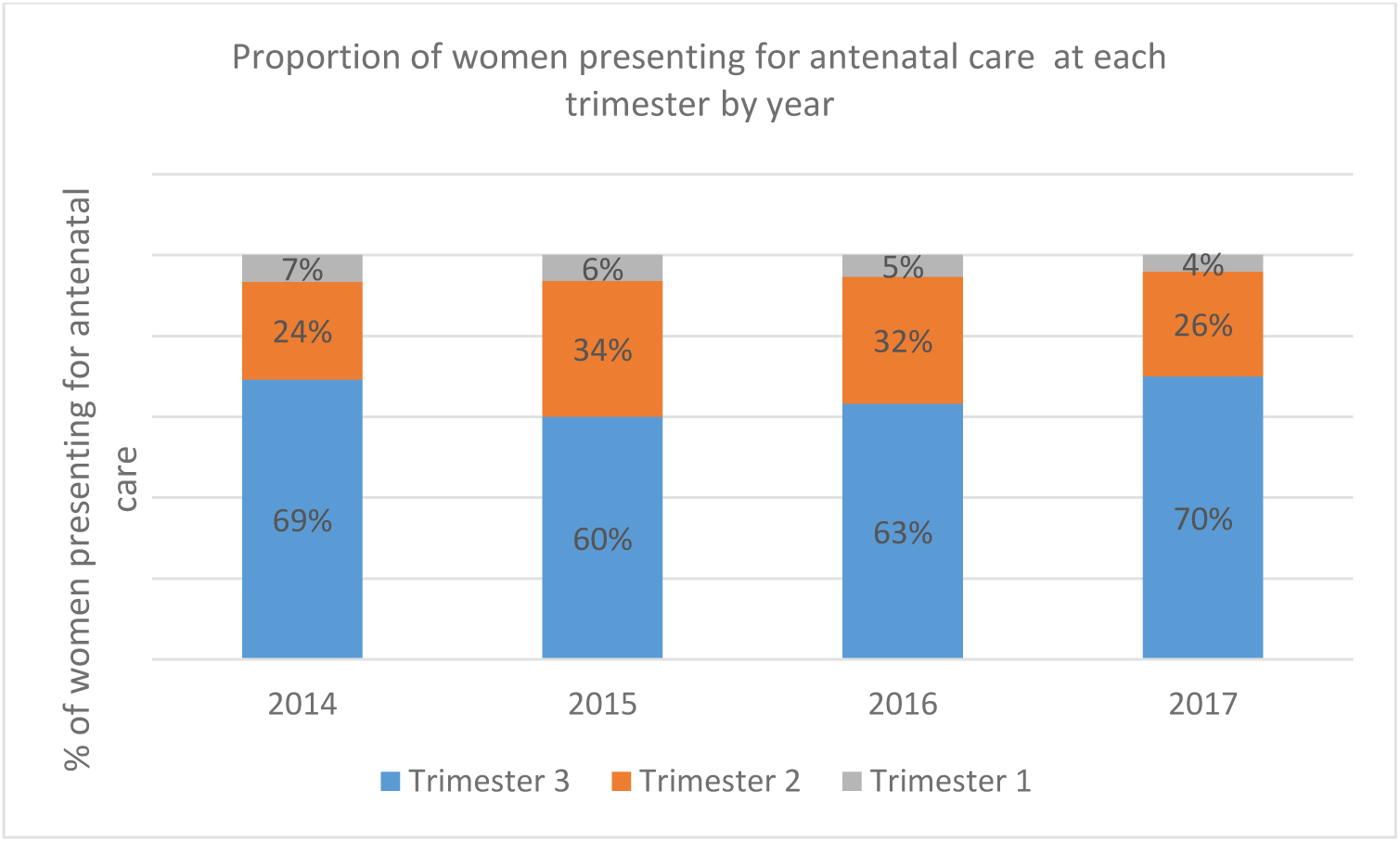
Timing of presentation into antenatal care by trimester (gestation age) over the years 2014-2017.

**Supplementary figure 2.**
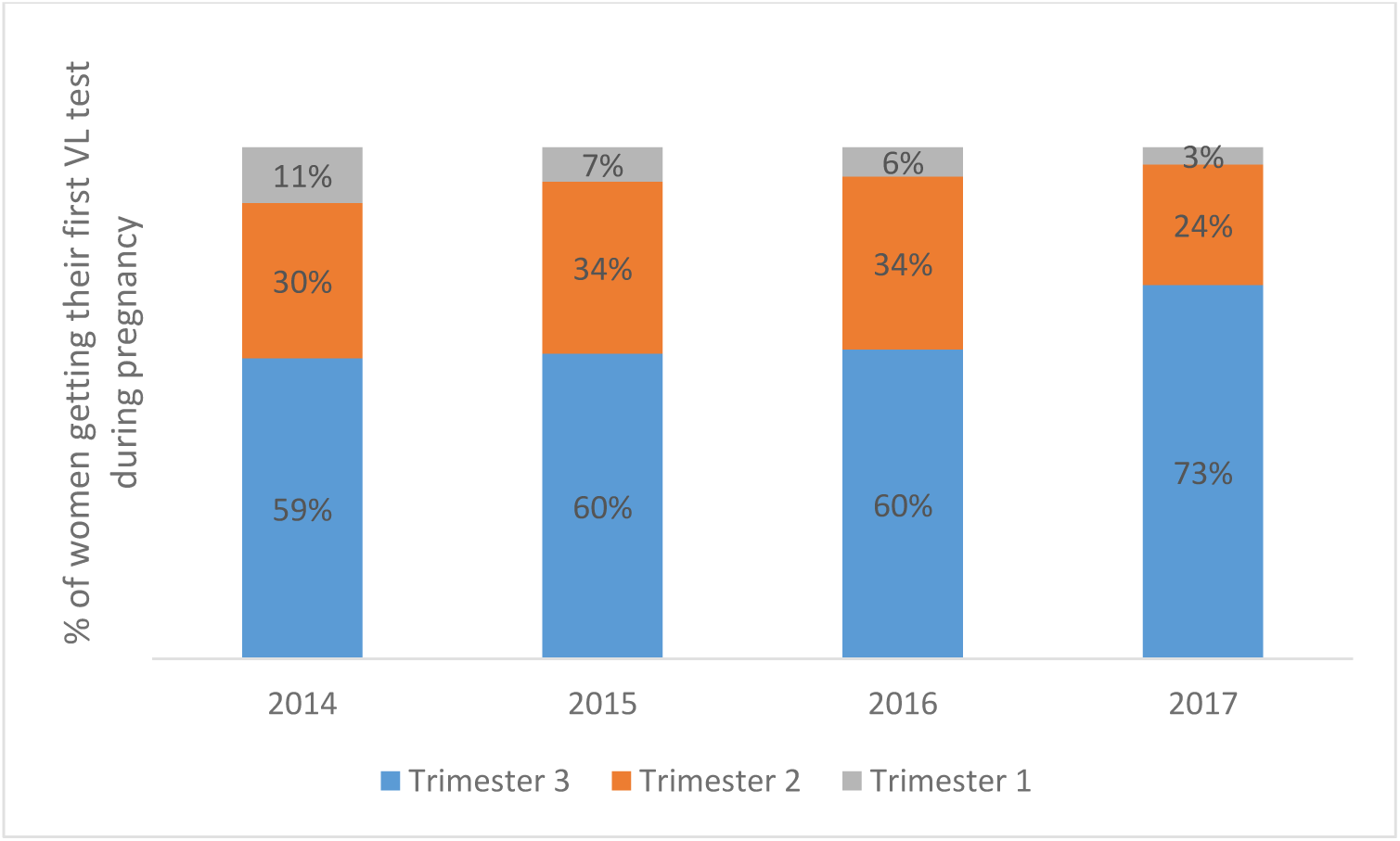
Timing of first VL by trimester (gestation age) over the years 2014-2017.

**Supplementary Figure 3.**
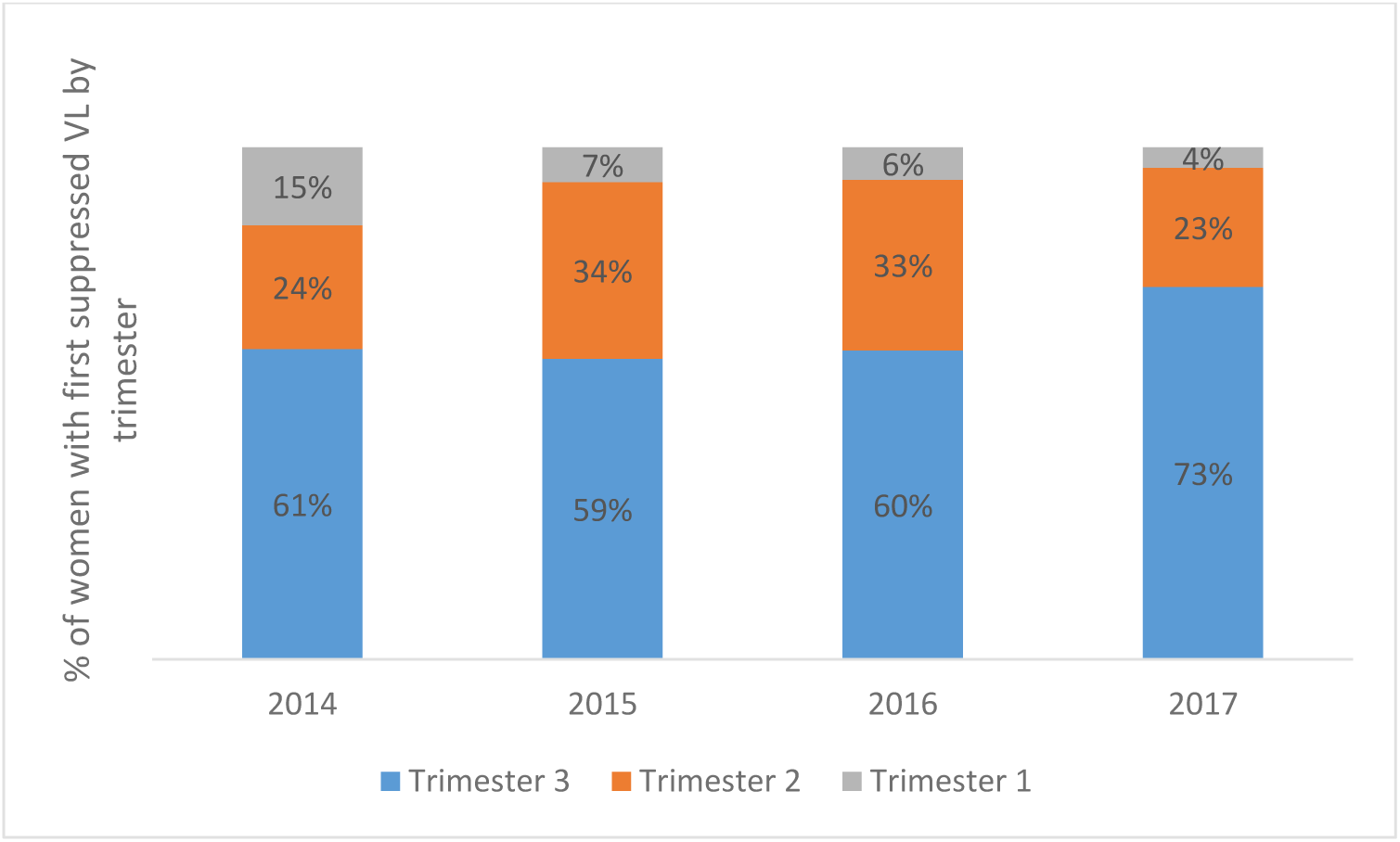
Timing of first suppressed VL results by trimester (gestation age) over the years 2014-2017.

## REFERENCES

[1] NDoH, “Guideline for the Prevention of Mother to Child Transmission of Communicable Infections South African National Department of Health,” Dep. Heal. Repub. South Africa, no. October, pp. 1–44, 2019.

[2] S. A. National Department of Health, “National consolidated guidelines for the prevention of mother-to-child transmission of HIV (PMTCT) and the management of HIV in children, adolescents and adults,” Pretoria, 2015. [Online]. Available: https://sahivsoc.org/Files/ARTGuidelines15052015.pdf

[3] National Department Of Health South Africa, “Guidelines for Maternity Care in South Africa. A Manual for Clinics, Community Health Centres and District Hospitals,” Pretoria, 2015. [Online]. Available: https://health-e.org.za/2015/11/17/guidelines-maternity-care-in-south-africa/

[4] World Health Organization, “New guidelines on antenatal care for a positive pregnancy experience,” 2016. https://www.who.int/news/item/07-11-2016-new-guidelines-on-antenatal-care-for-a-positive-pregnancy-experience

[5] R. Beauclair, G. Petro, and L. Myer, “The association between timing of initiation of antenatal care and stillbirths: A retrospective cohort study of pregnant women in Cape Town, South Africa,” BMC Pregnancy Childbirth, vol. 14, no. 1, pp. 1–10, 2014, doi: 10.1186/1471-2393-14-204.

[6] W. B. MacLeod et al., “Cohort profile: the South African National Health Laboratory Service (NHLS) National HIV Cohort.,” BMJ Open, vol. 12, no. 10, p. e066671, Oct. 2022, doi: 10.1136/bmjopen-2022-066671.

[7] K.-G. Technau et al., “12-month outcomes of HIV-infected infants identified at birth at one maternity site in Johannesburg, South Africa: an observational cohort study.,” lancet. HIV, vol. 5, no. 12, pp. e706–e714, Dec. 2018, doi: 10.1016/S2352-3018(18)30251-0.

[8] K.-G. Technau, L. Kuhn, A. Coovadia, S. Carmona, and G. Sherman, “Improving early identification of HIV-infected neonates with birth PCR testing in a large urban hospital in Johannesburg, South Africa: successes and challenges.,” J. Int. AIDS Soc., vol. 20, no. 1, p. 21436, Apr. 2017, doi: 10.7448/IAS.20.01/21436.

[9] A. C. of O. and Gynecologists, “Methods for estimating the due date,” Am. Coll. Obstet. Gynecol., vol. 129, no. e150–4, 2017, [Online]. Available: https://www.acog.org/clinical/clinical-guidance/committee-opinion/articles/2017/05/methods-for-estimating-the-due-date

[10] World Health Organization, “Consolidated Guidelines on the Use of Antiretroviral Drugs for Treating and Preventing HIV Infection: Recommendations for a Public Health Approach.,” Geneva, Switzerland, 2016. [Online]. Available: https://www.who.int/publications/i/item/9789241549684

[11] S. A. Woldesenbet et al., “Coverage of maternal viral load monitoring during pregnancy in South Africa: Results from the 2019 national Antenatal HIV Sentinel Survey.,” HIV Med., vol. 22, no. 9, pp. 805–815, Oct. 2021, doi: 10.1111/hiv.13126.

[12] F. Moyo, A. H. Mazanderani, T. Kufa, and G. G. Sherman, “Maternal HIV viral load testing during pregnancy and postpartum care in Gauteng Province, South Africa.,” S. Afr. Med. J., vol. 111, no. 5, pp. 469–473, Apr. 2021, doi: 10.7196/SAMJ.2021.v111i5.15240.

[13] C. C. C. Musanhu, K. C. Takarinda, J. Shea, I. Chitsike, and B. Eley, “Viral load testing among pregnant women living with HIV in Mutare district of Manicaland province, Zimbabwe,” AIDS Res. Ther., vol. 19, no. 1, pp. 1–11, 2022, doi: 10.1186/s12981-022-00480-1.

[14] M. Sandbulte, et al., “Maternal viral load monitoring: Coverage and clinical action at 4 Kenyan hospitals.,” PLoS One, vol. 15, no. 5, p. e0232358, 2020, doi: 10.1371/journal.pone.0232358.

[15] K.-G. Technau et al., “Timing of maternal HIV testing and uptake of prevention of mother-to-child transmission interventions among women and their infected infants in Johannesburg, South Africa.,” J. Acquir. Immune Defic. Syndr., vol. 65, no. 5, pp. e170–8, Apr. 2014, doi: 10.1097/QAI.0000000000000068.

[16] L. Parmley et al., “Antenatal care presentation and engagement in the context of sex work: exploring barriers to care for sex worker mothers in South Africa,” Reprod. Health, vol. 16, no. 1, p. 63, 2019, doi: 10.1186/s12978-019-0716-7.

[17] T. Kufa et al., “Point-of-care HIV maternal viral load and early infant diagnosis testing around time of delivery at tertiary obstetric units in South Africa: a prospective study of coverage, results return and turn-around times.,” J. Int. AIDS Soc., vol. 23, no. 4, p. e25487, Apr. 2020, doi: 10.1002/jia2.25487.

[18] M. R. Lamb et al., “High attrition before and after ART initiation among youth (15-24 years of age) enrolled in HIV care.,” AIDS, vol. 28, no. 4, pp. 559–568, Feb. 2014, doi: 10.1097/QAD.0000000000000054.

[19] E. Koech et al., “Characteristics and outcomes of HIV-infected youth and young adolescents enrolled in HIV care in Kenya,” AIDS, vol. 28, no. 18, 2014, [Online]. Available: https://journals.lww.com/aidsonline/fulltext/2014/11280/characteristics_and_outcomes_of_hiv_infected_youth.11.aspx

[20] W. A. Technau, K.-G. Maskew, M. Nattey, C. Hwang, C van Dongen, N., Ferreira, T., “Cohort Profile: Rahima Moosa Mother and Child Hospital Maternal HIV Cohort,” BMJ Open, vol. Manuscript, 2024.

